# Supporting Self-management Through eHealth - Exploring the Needs, Challenges and Solutions in General Practice: A Qualitative and Participatory Design Study

**DOI:** 10.1101/2024.05.17.24307464

**Authors:** Chris Djurtoft, Kristine Sørensen, Christian Odgaard, Morten Hoegh, Michael S. Rathleff, Simon K. Johansen

## Abstract

**Introduction:** Digital transformation and integration of eHealth solutions into chronic pain management faces significant challenges that have not yet been met. To realize the potential of eHealth solutions there is a need to understand the challenges, needs and care processes of eHealth into specific contexts and specific purposes. The objective of this study was to explore challenges, barriers, support needs, and visions experienced by patients and general practitioners (GPs) in the context of an eHealth solution designed for chronic pain management in general practice.

**Methods:** The study used action-research as a methodological framework. We conducted two future workshops involving eight patients living with chronic pain and seven GPs with clinical experience in managing chronic pain. Through case vignettes and inspiration cards, these workshops stimulated discussions and shared knowledge construction. Data were analysed using reflexive thematic analysis, separated by the groups, and were synthesized via a matrix analysis.

**Results:** The analysis revealed five content summary themes: Theme 1—patients’ experience of challenges in life with pain; Theme 2—challenges in treating patients with chronic pain; Theme 3—patients’ suggestions for the structure of the eHealth solution; Theme 4—GP’ suggestions for the structure of the eHealth solution; and Theme 5—differences and similarities: Visions for an eHealth solution. The analysis generated several touchpoints and tension within the patient-physician encounter.

**Conclusion:** In conclusion, these themes provide distinct narratives, offering valuable insights into the design objectives. Our study represents a significant advancement in developing personalized and innovative eHealth solutions for general practice, addressing key clinical challenges.

**Perspective:** Realizing the potential of eHealth solutions, these findings highlight both contrasting and shared viewpoints on design objectives, providing crucial insight into end-user perspectives for effective pain management. Additionally, the study underscores the importance of supported self-management and clinical communication in understanding each patient’s overall presentation within the healthcare system.

## Introduction

Chronic pain is a complex condition that can be challenging to live with and manage. This recognition describes the multifaceted impact of chronic pain on various aspects of an individual’s life; chronic pain is linked to diminished functionality, decreased social participation, and impaired workability ^18,48,49,72^. The utilization of healthcare services for chronic pain is noted to be frequent, placing a substantial burden on the healthcare system, particularly in general practice ^55,56^. General practitioners (GPs), who are often the first point of contact for individuals with chronic pain, find chronic pain to be one of the most challenging conditions to manage ^13,14^. The challenges are multifaceted, encompassing factors such as the complex nature of pain conditions, temporal constraints, diagnostic uncertainties, and a limited knowledge of effective pain and non-pharmacological management strategies. ^13,14,29,73^.

Novel solutions are needed to make best use of clinical resources ^8,29^. Electronic health (eHealth) solutions hold promise for bridging this gap since they are perceived as being affordable, adaptable to individual needs, and integrated into patients’ daily life ^7,8,24,69^. eHealth technologies, conceptually defined as: *“health services and information delivered or enhanced though the internet or related technologies”* ^21^ offer a wide range of possibilities for improving current and future pain management. This includes delivering health information, supporting self-management, monitoring symptoms, providing access to social/peer support, and facilitating on-demand contact with health resources ^30,34,39,52^. While eHealth solutions have the potential to significantly affect various aspects of healthcare, needs and design objectives regarding chronic pain management have not been thoroughly studied, highlighting a potential gap in research of eHealth solutions ^3,20,33^.

The digital health literature emphasizes that designing eHealth solutions for supported self-management of chronic conditions is complex, requiring insights into patients’ symptoms, perceived challenges, needs, stakeholders, care processes, and behaviours facilitating recovery ^22,28,40,41,51^. Stakeholder and user-participatory co-design is considered crucial for designing eHealth solutions ^75^; these methods are described as the gold standard for ensuring that eHealth solutions accommodate the domains of importance to all involved stakeholders ^3,35,77^. Nevertheless, diverging from these recommended standards, recent evidence concerning e-health for pain conditions highlights a noticeable deficiency in both end-user engagement and contextual integration ^33^. In pain management, treatment decisions are collaborative processes that often involve tensions, ‘opt-in’ and ‘opt-out’ on both sides during the consultation(s) ^30,44^. Therefore, real-world insights provided by patients living with chronic pain and GPs as lead users, drawing upon the generative activities and collaborations regarding the eHealth landscape is crucial for evidence-based design objectives ^47,75^.

The objective of this study is to explore the challenges, barriers, support needs, and visions experienced by patients and GPs in the context of an eHealth solution designed to support the management of chronic pain in general practice. Through two future workshops and qualitative analyses, our aims are: 1) identifying the experienced needs and challenges of end-users in managing chronic pain in general practice, and 2) extracting their perspectives and ‘visions’ on addressing these challenges using an eHealth solution. This will enable us to generate specific ‘design objectives’ for improving and implementing future eHealth solutions in general practice, meaningfully addressing the needs, gaps, and tensions of patients and GPs in pain management.

## Methods

### Study Design

We employed action-research (AR) as a methodological framework for structuring our investigation ^10^. We conducted two future workshops: one with patients experiencing chronic pain and another with GPs having experience in managing chronic pain in general practice. The workshops used case vignettes and inspiration card exercises to stimulate discussions and shared knowledge construction ^25,32^. Participant discussions emerging during each workshop were audio recorded, transcribed, and analysed separately via reflexive thematic analysis using NVivo (version 11; QSR International) ^9,64^. This study was reported in accordance with the guideline presented in the Consolidated criteria for reporting qualitative research (COREQ) ^71^. The study protocol was submitted for revisions to The North Denmark Region Committee on Health Research Ethics, which determined that the study was exempt from additional requirements based on Danish national guidelines.

### Research team and reflexivity

The research team, composed of five physiotherapists (CD, KS, CO, MSH, MSR), and one with a background in information science (SKJ). KS was the sole female member in the research team, which otherwise consisted of male members. Regular communication and collaboration among team members contributed to a cohesive research effort. The participants were contacted before the interview; otherwise, there was no relationship established prior to the interview, and there was no prior acquaintance between the interviewer and the participants. Reflexivity was integral, with each member acknowledging their positionality and biases, fostering ethical decision-making.

### Participants

To capture both patient and GP perspectives, this study included two populations. Population 1 consisted of patients experiencing chronic pain (duration >3 months), which could not be accounted for by a different diagnosis, as defined by the ICD 11 classification ^59^. Exclusions from this group included individuals with competing musculoskeletal or pain conditions, severe physical handicaps, and psychological issues that could impede participants’ ability to recall the pain experience and associated needs. Population 2 comprised GPs actively working in general practice with experience in managing patients experiencing chronic pain, demonstrating a willingness to participate in the study.

### Inclusion

For population 1, we recruited patients using social media posts aimed at individuals in the North Denmark Region. The posts contained a link to a form with questions related to the inclusion criteria, as well as contact information and consent forms. For population 2, we identified and recruited participants from this group through the Center for General Practice and Nord-KAP—the Quality Unit for General Practice’s clinician networks in the North Denmark Region. We contacted them via email and phone, providing study information, screening for eligibility, and extending invitations to participate. Those expressing interest and consenting to be contacted were subsequently informed about the project, their rights as participants, and the data treatment procedures.

### Future Workshops

The aim of future workshops is to create a safe environment for discussing real life issues, where participants can utilize their own experiences and perspectives formulate ideas and define novel solutions, which will facilitate desired changes in real life settings ^2^. We included the future workshop approach described by Apel ^2^ and Vidal ^76^ as a model for planning and conducting our interventions. A key feature of the future workshop relates to the methods reliance on co-construction of knowledge to empower participants to define and articulate future visions, though a three-step process focusing on critique, social fantasy, and implementation ^76^. To support participants transitions across the three workshop phases, we included inspiration card exercises and a case vignette to encourage participants dialogues and guide participants efforts towards formulating future visions (Table 1).

**Table 1.** Overview of the future workshop phases.

| Future workshop phases | Aim and methods. |
| --- | --- |
| Timeline and Phases | Brief explanation about the phases. |
| **Before**<br>Preparation | Organizers and facilitators agree on the theme, invited participants, methods, location, locales, rules, and timetables of the future workshop. |
| **The critique phase** | • Designed to draw out specific issues in question/producing a critical understanding of the problem.<br>• Collection of critique-points (posters, inspirational cards, brainstorm).<br>• Systemization/clustering on pinboards.<br>• Evaluation, condensation, and prioritizing. |
| **The fantasy phase (insights)** | • Imaginative productions (meditation, walks, work).<br>• Turn critique into opposites (good, bad) as starting points.<br>• Collecting ideas (brain writing, shot gunning).<br>• Analysis and elaboration of great ideas.<br>• Registration of ideas in idea book for later exploration. |
| **The implementation phase (visions)** | • Evaluate the registered ideas.<br>• Formulate best idea in concrete terms.<br>• Choose very best ideas.<br>• Action plan (solidify and implement) |
| **After**<br>Follow-up | Action plans are monitored, changes are performed, and new future workshops are planned to address challenges to implementation. |

### Pain Guide eLearning platform (Danish: SmerteVejleder)

Pain Guide eLearning (SmerteVejleder) is an online patient-education module with six sessions, each containing pain-related information, quizzes, and homework. We drew inspiration from Pain Guide eLearning for our workshops to prepare participants for active engagement. Our goal was to provide participants with foundational knowledge and awareness of eHealth solutions, enhancing their understanding and engagement. We intended to guide participants through a reflective process to identify their symptoms, concerns, and needs, fostering creativity and discussions before the workshops. Participants in both groups were granted access to the platform two weeks in advance and instructed to review the provided materials prior to the workshop.

### Case Vignettes

A case vignette (appendix 1) and inspiration card exercises were used to promote dialogue and shared construction of knowledge during each workshop ^25,32^. The shared learning activities of the workshop was anchored in a case vignette, which is designed specifically to outline the salient features of a case: *‘a patient with long-standing chronic pain in general practice’*, while providing participants with a starting point for discussions and the card exercises. The case was created in collaboration with GPs and patients experiencing chronic pain, containing useful and unusable information.

### Inspiration Cards

A card exercise for inspiration was developed with the aim of fostering dialogue and collaborative creation, while also providing guidance to participants throughout the three phases of future workshops (appendix 2). To ensure the themes’ relevance and comprehensibility, the inspiration cards underwent testing and iterations following a similar pattern as the case vignettes.

### Other Artifacts

Each workshop was equipped with additional materials. PowerPoint presentations was employed by the workshop facilitators to lead the workshop activities. All participants used post-its (red, yellow, and green), pens and markers for writing notes and making up new cards for the inspiration-card games, as new topics may emerge through the dialogues. Whiteboards, flipboards and poster-size sheets of paper was used by the facilitator (SKJ) to guide plenum discussion of visions/insights and visualize and collect findings. These tools were intended to facilitate the brainstorming process, enabling participants to generate ideas collaboratively. The materials enabled participants to organize emerging themes, allowing participants to visually establish conceptual relationships.

### Setting and Procedure

The workshops were conducted within the Center for General Practice at Aalborg University. Based on the recommendations from Kanstrup & Berthelsen, we aimed for 8-16 participants (patients and GP’s) ^31^. This number of participants will ensure a meaningful dialogue and enable co-construction between the stakeholders during the workshops. Apart from the participants, the workshop involved the participation of two researchers: one acting as a facilitator (SKJ) and one taking on the role as observer and co-facilitator. To promote a safe environment for discussions and avoid asymmetric power relationships, the study populations were kept separate during each workshop. To provide participants with an equal opportunity to be heard during interventions, participants were divided into workgroups of three to four participants ^31^. Each of the workshops was initiated by a short 15-minute presentation describing ‘Pain Guide eLearning platform’ and the aim of the project. This was followed by three phases of 45-minute sessions inspired by the three phases of the future workshop (critique, fantasy, and implementation) ^28,31^. The participants were asked to use the inspiration cards and case vignette to discuss and share their perspectives and experiences on living with chronic pain during the phases (appendix 3) ^28,31^.

After each of the three sessions, a short plenary was conducted where participants shared perspectives and received feedback from the other groups. All ideas and possible solutions for improving the eHealth program were noted on a whiteboard through plenary discussions. At the end, all participants were debriefed, informed about the rights as informants and asked to fill in consent forms and reimbursement sheets.

### Data Collection

We gathered baseline demographics from the two participating groups using REDCap (Research Electronic Data Capture) ^26^. Each participant received a link and filled out the questionnaire on their smartphone. The format remained consistent across the participants and tailored to suit the specific characteristics of each target group. The participant’s dialogue within the workgroups were recorded using an Olympus WS-553 dictaphone. Subsequently, all data was uploaded to a secure server hosted at Aalborg University. Two participants choose not to answer some baseline questions (referred to as ‘preferred not to answer’).

### Analysis

The analysis was conducted in two separate sprints, before being combined for the final interpretation, synthesized in a matrix analysis ^4,9^. This included 1) read-throughs of the whole dataset and 2) thematic coding of meaningful units, 3) generation of main- and sub-themes, 4) selection, and 5) condensation, refinement, and interpretation ^9^. Finally, the data was merged through constructions of a framework to generate intertextual overlaps/divergencies and extract analytical thematic categories.

The qualitative data collected during the two future workshops were transcribed and analysed in accordance with Braun and Clarke reflexive thematic analysis by the lead researchers (CD and SKJ) and two research assistants (KS and CO) ^9^. The data sets from each workshop were transcribed for meaning retention, as described by Kvale and Brinkmann, using Microsoft Word ^37^. Transcribed data were analysed in parallel through a four-stage process including familiarization, coding, and generating themes, condensation and refinement, and synthesis into a shared narrative according with Braun and Clarks six phases ^9^. NVivo (version 11; QSR International) coding software was used for the coding and organization of themes, and coding lists were created and maintained by all coders (KS, and CO) during each individual analysis ^64^. KS and CO conducted the initial coding and thematization independently of each other; subsequently, data were double coded to ensure concise definitions for each code and theme. Generated themes were refined through iterative cycles of horizontal readings and condensation, and related subthemes were merged. To ensure coding integrity, we used stakeholder checks, and coding list entries were discussed as the analysis progressed; this were employed via multiple group meetings of all involved parties. We employed mind-maps to consider the thematic relationships developed during each individual analysis. Thematic overlaps, divergencies, and relationships within the individual analysis were discussed among CD, SKJ, KS and CO, until consensus was reached. Lastly, the findings from each workshop were synthesized in a matrix analysis to explore how the eHealth app could support patients and GPs in treatment ^4^. The insights uncovered during each thematic analysis were organized in a matrix to map collaborative tensions; this informed the design objectives, and future implications of the eHealth design ^4,9^. The matrix analysis helped us understand areas of needs, potential agreements/disagreements, sources of tension, and challenges. This facilitated the articulation of a set of design principles for designing eHealth solution for supporting the treatment of patients experiencing chronic pain in general practice.

CD and SKJ was responsible for the final abstraction and presentation of the findings in the five storybook themes, which outlined the narrative (Results section). All involved parties were involved in the analysis. CD, SKJ, KS, and CO approved the first draft of the storybook themes, narrative, and matrix analysis before the analysis was concluded and validated by all authors.

## Results

### Inclusion of Participants

The inclusion produced eight patients experiencing chronic pain (workshop 1), and seven GPs (workshop 2) (Figure 1).

**Figure 1.**
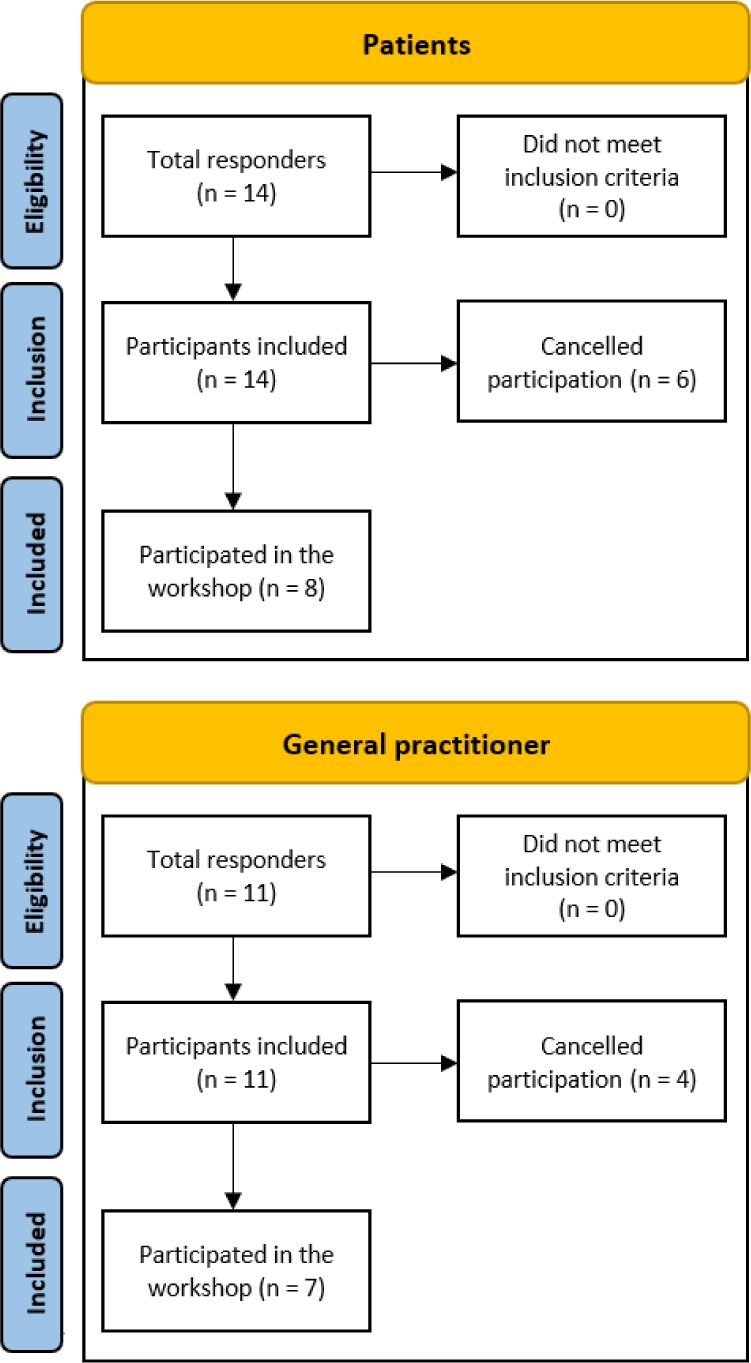
A flowchart providing an overview of the two participating groups.

All patients were women (n=8), with an age range of 32-76 years. Most GPs were women (n=5), having an age range of 29-46 years (Table 2).

**Table 2.** Participants demographic data.

| **Patients (n=8)** |  |
| --- | --- |
| **Age** median (range) | 44 (32-76) |
| **Gender** | Female: n=8 |
| **Nationality** | Danish: n=8 |
| **Job situation** | Full time: n=4<br>Part time: n=2<br>Retired: n=1<br>Preferred not to answer: n=1 |
| **Pain duration (years)** median (range) | 22 (3-44) |
| **Pain in body parts/area** | Neck: n=4<br>Back: n=4<br>Abdominal: n=1<br>Shoulder: n=3<br>Arm: n=2<br>Wrist: n=2<br>Hand: n=1<br>Hip: n=2<br>Pelvic: n=3<br>Thigh: n=1<br>Calf: n=1<br>Foot: n=1<br>Headache: n=1<br>Fibromyalgia: n=3<br>Preferred not to answer: n=1 |
| **Pain medication** n= (%) | No: n=1 (12,5%)<br>Rarely: n= 0 (0,0%)<br>Once a month: n= 0 (0,0%)<br>Once a week: n= 0 (0,0%)<br>More than once a week: n=1 (12,5%)<br>Daily: n=4 (50,0%)<br>Preferred not to answer n=2 (25%) |
| **General practitioners (n=7)** |  |
| **Age** median (range) | 35 (29-46) |
| **Gender** | Female: n=5<br>Male: n=2 |
| **Nationality** | Danish: n=7 |
| **Specialty** | General practitioner: n=7 |

### Results of the Data Analysis

The coding and analysis of data from Workshops 1 and 2 generated five themes which informed a narrative of five content summary themes (Table 3). The five content summary themes were: Theme 1: patients’ experience of challenges in life with pain; Theme 2: challenges in treating patients with chronic pain; Theme 3: patients’ suggestions for the structure of the eHealth solution; Theme 4: GP’ suggestions for the structure of the eHealth solution; and Theme 5: Differences and Similarities between Patients and Doctors: Visions for an eHealth solution.

**Table 3.**
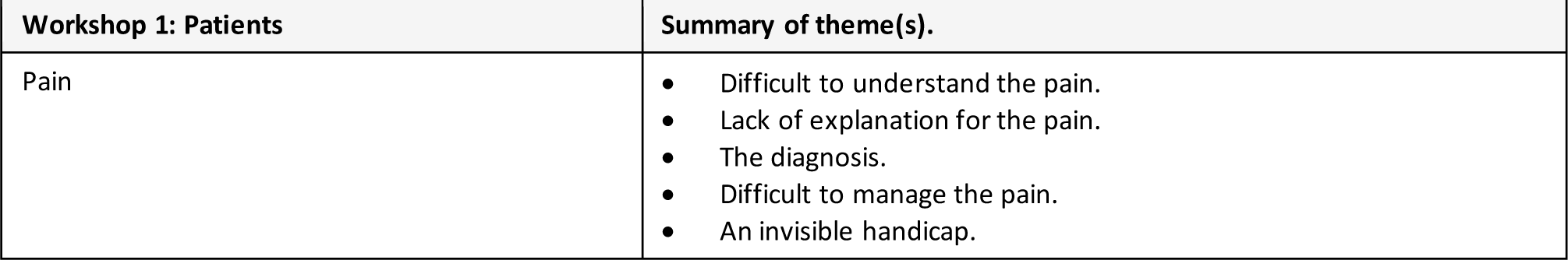
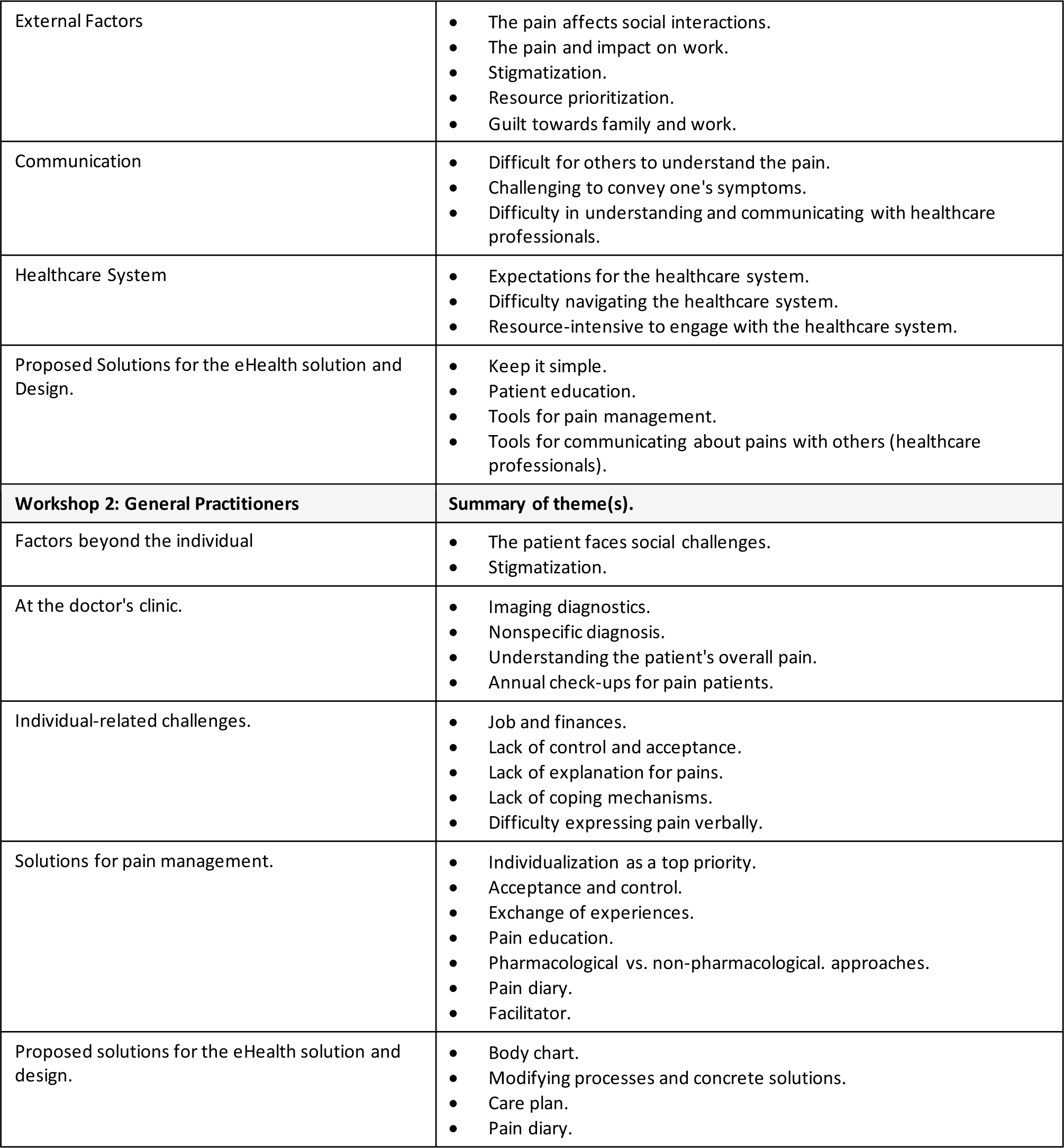
Content summary the five themes.

| **Workshop 1: Patients** | **Summary of theme(s).** |
| --- | --- |
| Pain | • Difficult to understand the pain.<br>• Lack of explanation for the pain.<br>• The diagnosis.<br>• Difficult to manage the pain.<br>• An invisible handicap. |
| External Factors | • The pain affects social interactions.<br>• The pain and impact on work.<br>• Stigmatization.<br>• Resource prioritization.<br>• Guilt towards family and work. |
| Communication | • Difficult for others to understand the pain.<br>• Challenging to convey one's symptoms.<br>• Difficulty in understanding and communicating with healthcare professionals. |
| Healthcare System | • Expectations for the healthcare system.<br>• Difficulty navigating the healthcare system.<br>• Resource-intensive to engage with the healthcare system. |
| Proposed Solutions for the eHealth solution and Design. | • Keep it simple.<br>• Patient education.<br>• Tools for pain management.<br>• Tools for communicating about pains with others (healthcare professionals). |
| **Workshop 2: General Practitioners** | **Summary of theme(s).** |
| Factors beyond the individual | • The patient faces social challenges.<br>• Stigmatization. |
| At the doctor's clinic. | • Imaging diagnostics.<br>• Nonspecific diagnosis.<br>• Understanding the patient's overall pain.<br>• Annual check-ups for pain patients. |
| Individual-related challenges. | • Job and finances.<br>• Lack of control and acceptance.<br>• Lack of explanation for pains.<br>• Lack of coping mechanisms.<br>• Difficulty expressing pain verbally. |
| Solutions for pain management. | • Individualization as a top priority.<br>• Acceptance and control.<br>• Exchange of experiences.<br>• Pain education.<br>• Pharmacological vs. non-pharmacological approaches.<br>• Pain diary.<br>• Facilitator. |
| Proposed solutions for the eHealth solution and design. | • Body chart.<br>• Modifying processes and concrete solutions.<br>• Care plan.<br>• Pain diary. |

The themes yielded distinct narratives providing valuable insights into the participants’ visions for the design objectives. Themes 1 and 3 primarily highlight patients’ perspectives, while Themes 2 and 4 emphasize the perspectives of GPs. Theme 5 stands as a separate theme, delving into the differences and similarities in the visions of patients and GPs for an eHealth solution. This theme offers profound insights into the complexities that both impede and facilitate the design process, underscoring the multifaceted nature of the two groups’ perspectives and emphasizing the potential for innovation and improvement in eHealth solutions. The matrix, in turn, served as a foundation for specific design objectives, serving as a foundational framework for specific design objectives. It delineates the dynamics at play within the design, aiming to foster shared perspectives and navigate tensions regarding the eHealth treatment of pain.

### Overview of Themes

The generated themes provided an indication that there were multiple perspectives at play regarding design objectives of the eHealth solution, both between groups and among individuals within the groups. The challenges and opportunities were broad ranging; this was expressed through both agreements and disagreements, highlighting potential tensions across the two participating groups.

#### Theme 1— Patients’ experience of challenges in life with pain

The first theme highlighted the challenges that patients with pain may experience in their daily lives. The patients described finding it difficult to understand their pain; they conveyed a profound sense of frustration stemming from the absence of a clear explanation for why their pain developed and a corresponding deficit in guidance on effective management strategies. The patients described that uncertainty about the pain could lead to concerns about its severity and duration and several patients highlighted the impact of a diagnosis, which could have both positive and negative implications. Some saw a diagnosis as providing them with an explanation and, in turn, inner peace and validation, while others viewed it as potentially offering access to financial benefits (such as early retirement) or acceptance from others. On the other hand, some patients had experienced receiving a diagnosis as being associated with loss and grief. It was described how a diagnosis could bring peace by validating their condition.

> *Patient D4: “Yes, I know from myself, when I got a diagnosis. It gives me peace within myself because then I was right. It’s not just something I make up. I’m good enough. And it’s not just whining. You find some peace within yourself, I think. And that can also be the positive aspect of getting a diagnosis.”*

The patients described how it was challenging for others to understand the implications and impact of their condition because it is not visible. They experienced a lack of understanding from family, friends, or co-workers, which made it difficult for the patients to explain their condition to them. The patients also found it challenging to communicate with healthcare professionals and to navigate the healthcare system. Some found it challenging to understand medical language/terms, while others struggled to express their symptoms. In addition to this, some patients experienced sleep problems or memory, concentration difficulties, and cognitive challenges, which negatively affected their ability to communicate with healthcare professionals:

> *Patient D3: “Yes, you can also sit with the doctor like that. Don’t you know that feeling when you’re sitting there going through the things you want to say before you go in, but then when you get in there, you’re just blank.”*

Patients expected GPs to provide a thorough evaluation and refer them to the appropriate treatment. However, they described being referred to various locations within the healthcare system without a clear understanding of their condition and what might be effective for them. Some patients experienced guilt when taking time off work to see the GP. Furthermore, patients felt burdened when their GP could not address their challenges despite repeated attempts. They described accessing healthcare as a resource-consuming process, leading to frustration with repetitive visits to GPs:

> *Patient D1: “Yes, it’s like running your head into a wall. You run in circles. Because you can go to the GP many times without getting any wiser. And it’s such a negative spiral that you get into. One question gives rise to ten others.”*

A recurring theme was the feeling of guilt that the patients experienced towards their friends, family, and work. The patients desired to be able to work and have the energy to spend time with their families. They described feelings of stigmatization and prejudice when on sick leave, rather than receiving understanding for their situation. Moreover, they found it challenging to ask their family for help. One patient described the guilt they felt in relation to their work:

> *Patient D5: “But the guilt comes anyway when I have to call in sick again. And then the others say, ‘Wow, you’re sick so often. You’re not really sick, are you?’”*

Patients expressed a desire for *”help to help themselves”* to be the *”captain of their own ship.”* At times, they needed someone else to take charge when they lacked the energy. Patients explained that the need for pain management tools varied individually, and the effectiveness of these tools depended on their personal characteristics and the duration of their pain. It was described that there was a need for different tools at different stages:

> *Patient D2: “I’ve been a pain patient for over 20 years, and among all those I’ve talked to, it seems that in the first many years, you have to find your ways, and at that time, you need something different than later in your process, where you have gotten to know yourself. And then you might also be more open to various solutions and treatment options. In the beginning, it’s okay to go to a chiropractor or a physiotherapist, but your own active part is not very significant because you’re going through a grieving process, and then you just can’t handle it.”*

#### Theme 2—GP’ experiences of challenges in treating patients experiencing chronic pain

This theme described the challenges that physicians perceived in treating patients experiencing chronic pain. GPs believed that there were challenges associated with giving patients a nonspecific diagnosis. They thought it would lead to stigmatisation of the patients and that the patients would experience a lack of acceptance and understanding of their pain. The GPs also described that patients often lacked coping strategies to manage their pain. It was described how GPs believed that a non-specific diagnosis could be challenging for patients experiencing chronic pain to understand:

> *GP D11: “(…) The stigma that can be from the surroundings when she can’t put words on it. So, when she has a hard time relating to it herself, then the surroundings also have a hard time relating to it. And it stems from when she comes to the doctor and is told it’s nonspecific lower back pain. Does it mean nothing? Is it nothing? Is it not serious? Or is it something? So, it’s about acceptance and recognition of what it means to have pain.”*

In connection with this, GPs understood that it becomes difficult for patients to explain their condition, which can have an impact on various aspects of the patient’s life. These aspects encompass employment, finances, social life, and negative emotions/thoughts:

> *GP D12: “Yes, and no matter what she does, she is influenced by her difficulty in understanding what is happening to her. She can’t put it into words and explain it to others. And she may feel shame or condemnation from others. And it spreads to all spheres of her life. So, we talked about how her quality of life is difficult to find because no matter what she does, she struggles.”*

To uncover how the above processes can influence patients’ pain experiences, GPs described the importance of considering pain from a biopsychosocial perspective, where all aspects of the patient’s life are explored. This includes potential issues like divorce, concerns about their children, or work-related worries. The GPs described that the patient’s pain could be better understood by considering the concept of “total pain.” (i.e., all possible dimensions of their pain). There was a broad understanding that pain is a complex condition, and GPs acknowledged that comprehending the entirety of it, such as how pain could affect the patient’s overall well-being on multiple levels, often extending beyond the pain itself, could be challenging:

> *GP D9:”Because it’s all sorts of other things that hurt. And that’s where we sometimes find ourselves in a situation where we haven’t really understood what the biggest problem is for the patient, but it becomes back pain or other pains that become the tangible expression that we’re not doing well.”*

GPs experience this challenge as a dilemma between not always knowing the cause of the pain but having to accept that “the patient has the pain the patient has,” and the stigma they believe it can cause to explain pain as a “*sociomental thing”*. Additionally, GPs also acknowledge that there may be biological causes of pain that are not yet known.

#### Theme 3— Patients’ Suggestions for the development of the eHealth solution

One of the main suggestions from patients for designing the eHealth solution was to “keep it simple”. They desired short and precise explanations in plain, non-medical language. They proposed the inclusion of a toolbox with tools for acquiring knowledge about pain, as well as managing and communicating pain. Patients recommended using headings or categories to make it easier to follow and review previously read information. The eHealth solution should allow each individual patient to choose the sections that are relevant to them and opt out those options, which are not aligned with what matters in their individual life.

> *Patient D2: “Yes, the toolbox (common denominator) can include many different tools and models. Pain scale. How long have you had your pain? How do you become the captain of your own ship? We all have our preferences. Of course, we do, because we are all different.”*

The patients emphasized that education about pain was an important component of the eHealth solution, providing information about what pain is and how to manage it. They recommended incorporating features such as, text or video content about pain. Additionally, they suggested including positive patient stories from other pain sufferers to offer hope, learning opportunities, and a sense of not being alone.

> *Patient D4: “Because there are others in the same situation. You can feel like you’re the only one experiencing it this way. Even though two stories are not the same, you can almost always find something that fits in. And then you can also see that you can gain something from the situation you’re in. Look at it in a positive way. Find the positives in negatives.”*

The patients desired tools for self-management, or *“help for self-help,”* to address the individual challenges they were facing. Some of the most frequently mentioned tools included a pain diary for communication and overview, a checklist for cognitive and emotional challenges, tools for goal setting, resource management, and exercise guidance:

> *Patient D5: “We talked a lot about this toolbox. And it pretty much covers everything because everything is a toolbox. And in there, we should have the options to pick the things that suit each of us. It’s hard to create something that fits all of us here in this room. So it’s important that we have the ability to pick from different shelves. For example, a checklist that can help one remember what to ask the doctor. Or a pain diary. Or mindfulness. Relaxation. Different apps one can use. Exercises to handle overthinking.”*

The patients suggested tools for communicating about their pain, such as a chat feature and functions to connect with a healthcare professional with expertise in pain, relatives, or other individuals experiencing pain. Additionally, the patients mentioned the need for a pain dictionary comprising definitions and descriptions to facilitate their understanding of medical jargon. Such a resource would effectively assist them in communicating about their pain to others:

> *Patient D2: “Yes, and I think that’s part of the toolbox, speaking the same language. Maybe you can also learn the terminology that professionals use regarding pain, pain patients, etc., right in there.”*

In terms of tailoring the eHealth solution to the individual, some patients recommended the inclusion of a user profile or questionnaire to be completed at the outset. The purpose of the user profile was to enable patients to keep track of what they had already read or watched, adjust the amount of information, or (de)select the most relevant tools. Additionally, they wanted the option to choose whether to navigate through the eHealth solution sequentially or handpick the content they found most relevant to their needs:

> *Patient D5: “Yeah, it’s again about where you are in your journey. For me, it’s important that it’s intuitive and easy to use, so you can select the things you’re curious about and click into each individual toolbox. It could be to-do lists or something similar. And there might be others who want to learn about pain, so they click on that box.”*

#### Theme 4— GP’ Suggestions for the development of the eHealth solution

The central theme from the GPs’ recommendations for eHealth solution development focused on individualisation. This theme underscores the importance of tailoring the eHealth solution’s content to meet the unique needs and preferences of each patient. GPs emphasised the need to offer a range of options within the eHealth solution, encompassing various learning styles and levels of detail. It was highlighted as essential to provide patients with various options within the eHealth solution, such as a simple version, a detailed version, and video materials, allowing each patient to choose based on their preferred learning style. By allowing patients to select from simple and detailed versions, as well as providing video materials, the eHealth solution may effectively cater to the individualised requirements of each user, making it personalised and adaptable for patients to engage with the eHealth solution in a way that best suits their learning and coping style.

The GPs described the individualisation through the personalised care processes. These care processes were envisioned to encompass treatment goals and annual check-ups, providing an overview of what should be addressed at the next medical consultation. Inside the eHealth solution, patients should have the capability to manage and comprehend their pain through fundamental education in physiology, such as the autonomic nervous system. Key elements of these care processes within the eHealth solution included a pain diary, patient education regarding pharmacological adverse events, and resource management. Additionally, involving relatives in the care processes and treatment was emphasised, as it was seen to create acceptance and understanding for the person experiencing pain, particularly in their social context. Concrete tools to manage pain were suggested, such as mindfulness, breathing exercises, and physical training, which patients could find inspiration for within the eHealth solution. Moreover, GPs emphasised to have the ability to *”insert links into their care processes”* regarding what the patient should work on at home, further personalising the eHealth solution’s content and care recommendations:

> *GP D11: “We call it the care plan for chronic pain sufferers. And it includes all these different things here. Education. Pain diary. We have coping strategies in various forms. Pharmacological side effects. And then, we have a little something for the relatives as well.”*

A theme among GPs was the inclusion of modifying processes and concrete solutions. This theme manifested in a specific solution where the patient initiates the process by completing a task, such as a body chart or describing their pain. By doing so, the patient ends up with a comprehensive self-assessment (i.e., a personalised complete picture of themselves). Subsequently, the patient would analyse the modifying processes contributing to their pain and identify which one needs attention. Working on the identified modifying processes should involve the possibility of accessing relevant information and guidance through the eHealth solution. The primary intention behind this approach is to shift the patient’s focus from pain towards controllable aspects of living with pain. Several GPs envisioned how this could generate feelings of empowerment and control for the patient, while also enabling doctors to assess progress, processes, and outcomes in treatment. GPs believe that they or another healthcare professional can serve as a supportive figure in helping the patient concentrate on the appropriate modifying processes. A sub-topic in this context involved engaging relatives in a type of game or activity to redirect their focus from pain and promote acceptance of the patient’s pain. According to the group, this model would result in empowered patients throughout their treatment:

> *GP D12: “(…) We should empower our patients. Make them competent. Create strong patients in their treatment. And this ties in with individualization. (…) Instead of focusing on where the pain is located, we should remove the focus and move it to these modifying factors. It’s like a picture (draws circles around the patient to indicate the different modifying factors) and then, the patient has a guide who says, “if we move this modifying factor now and then it turns green” – is it easier for you to handle your pain?”.*

A theme among the GPs highlighted the need for a guide, which could take the form of a coordinator, or an annual check-up structured in general practice. Within this theme, GPs engaged in discussions about whether to refer to them as “annual check-ups” or “follow-ups” and whether to designate them as “care processes” or “care plans”. Furthermore, the GPs expressed a belief that follow-ups and coordination could be managed by various healthcare professionals, and possibly other people living with pain:

> *GP D10: “I certainly think that there should be some form of follow-up. A facilitator. A process that is clear from the start. A pain guide, a professional. It can be a doctor, nurse, physiotherapist, or similar. Someone who practices and takes care of these patients. Someone who has the responsibility and can help and is trained for it. I think that’s relevant and necessary for it to work, so that they don’t fall through the cracks”.*

#### Theme 5— Differences and Similarities between Patients and Doctors: Visions for an eHealth solution

The two participant groups offered different perspectives and highlighted both distinct challenges and similar solutions regarding what they considered as important eHealth design objectives in the treatment of chronic pain. GPs described how they sometimes could not provide patients with a sufficient explanation for their pain. They believed it was crucial to help patients understand that pain is a multidimensional experience. However, they also acknowledged that this posed a significant challenge as it increased complexity. Contrary to this, patients highlighted the importance of receiving a diagnosis and an explanation for their pain; they emphasized repetitive feelings of frustration as they were unable to obtain the desired explanation from their GPs. Additionally, patients frequently described the experience of visiting a GP multiple times without finding relief. They found it unclear what treatment options were available when the GP was not able to provide effective management:

> *Patient D7: “Yes, well, GPs today are also…Well, general practice – it’s broad. It’s difficult because they can’t know everything, and that’s fair enough. But it’s true – what options do they have to refer us? Because nonspecific back pain? Seriously? “We could have come up with that ourselves.”*

Patients experienced difficulties in communicating their pain to healthcare professionals, which was not articulated in the same way by the GPs. Patients found it challenging to explain their pain to GPs, whereas GPs described that they did not always fully understand the patient’s problem(s). As a solution to this issue, patients suggested maintaining a pain diary or using tools within the eHealth solution to express their symptoms. The recommended tools included pain education or a dictionary with examples of how to describe their symptoms and other domains of difficulties in their life. The pain diary would help them organize their problems for themselves, facilitate communication of their symptoms to the GP, and enhance learning about themselves. GPs proposed that patients create a ‘self-portrait’ through the eHealth solution, allowing it to display a realistic image of the patient with symptoms and modifying processes of change that could be personalised. This information would be automatically organized into sections visible only to the GP, enabling them to gain an overview of all aspects of the patient’s problem:

> *Patient D2: “Yes, we also talked about having a common language between healthcare professionals and us patients. Maybe there could be a small library in there, explaining what I mean when I say I have back pain, so it means this. Like how the doctor could write what he/she means when using specific terminology. Then maybe we could have a meeting at some point.”*

Both patients and GPs described the concept of self-management as important. Patients expressed challenges with problems they faced in their daily life, emphasizing the need for tools to support them. They desired the ability to address these issues independently, but they pointed out that they lacked knowledge about how to effectively manage these problems and where to seek help when needed. Therefore, patients frequently mentioned the importance of pain education, encompassing education in personalised pain management. Patients stressed the variability of their needs, underlining that the tools in the eHealth solution should be individualised to accommodate these variations. On the other hand, GPs described their focus on diagnosing patients and gaining a comprehensive overview of the patient’s entire situation, including modifying processes. They positioned themselves in a supportive role, with a greater emphasis on non-pharmacological treatments for patients. GPs explained that their goal was to empower patients so that they could handle and self-manage their pain through pain education and the personalised management of various processes and domains in their lives. They believed that this supportive personalised approach would provide patients with a sense of control and understanding of their pain, ultimately helping them manage their life living with pain:

> *GP D12: “It’s individualization. Regardless of which treatments and strategies we need to use. Until now, we’ve talked more generally about one platform for everyone, but as individuals, we have differences in our mental abilities, etiology, and reinforcing factors. For example, this patient has been very isolated and mentions something about sleep, whereas other patients may have other modifying factors that require different management.”*

While patients expressed their desire for self-management, they also acknowledged that there are times when they require guidance. GPs considered this perspective important and proposed the idea of having a contact person or coordinator to follow up with the patient. The objective of this role is to ensure that “the patient does not get lost” and “feels supported”. Such follow-ups could occur during an annual check-up or video consultations. Patients themselves suggested the inclusion of a chat function within the eHealth solution, allowing them to connect with a healthcare professional or another person having the role as a supervisor (i.e., a person living with pain). This approach is intended to offer an online channel for seeking guidance, support and self-management solutions when needed:

> *Patient D7: “Having someone who knows a little more or is wiser than us and can guide us in the right direction. A bit like how our children say they also need an adult from time to time. We need someone who can guide us.”*

### Results of the Matrix Analysis

The matrix analysis generated several touchpoints and tension which emerged within the patient-physician encounter, and created insights into how a digital solution could help bridge the communicative gap between GPs and patients. A key finding related to how GPs and patients took on different roles within the treatment situation. The patients described how they contacted their GP with the intent of getting a diagnosis, and advice for how to master their everyday pain autonomously, but also how they sometimes needed GPs to take charge and make decisions for them. While patients described how not being met with understanding by GPs was frustrating and led to worries and tensions, both GPs and patients described how patients would often struggle to remember and articulate developments in their pain and how this exacerbated the communication issues and made collaborations challenging.

On the contrary, the GPs described how they saw themselves in a supportive role during treatments, but also expressed the difficulties in this supportive role. While acknowledging the impact of pain on various aspects and domains of patients’ lives and expressing a desire to assist with issues such as employment or stigma, the GPs struggled to attain a comprehensive understanding of the patients’ situations. This lack of complete information caused tensions, as GPs felt they lacked sufficient insight to offer quality advice. Furthermore, the GPs highlighted how providing a diagnosis could be seen as a validation for having pain and help patients to accept their condition. Some GPs were concerned about how giving patients a non-specific diagnosis; they believed it would influence patients’ management of their negatively and this was something GPs wanted to avoid. Finally, the GPs used referrals as a way of gaining a better understanding of the patients’ symptoms, patients felt that being sent off to a specialist as a negation of their experience.

The findings from the matrix analysis generated several core features for a digital solution which could be included to alleviate tensions and help patients to bridge the communicative gaps during treatments (Figure 2).

**Figure 2.**
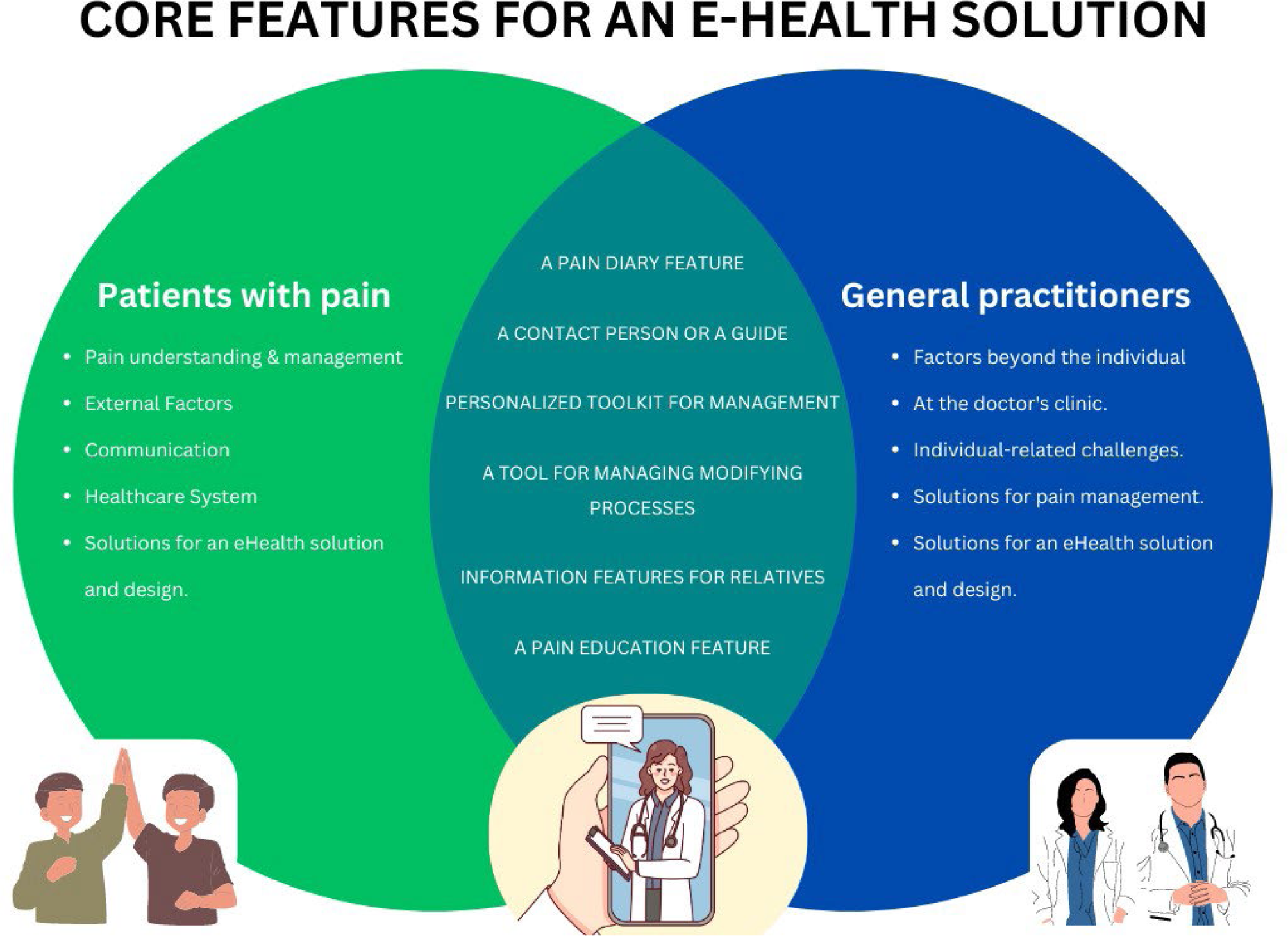
Venn diagram. The Venn diagram captures the collective vision of patients and GPs, illustrating the shared and distinct features proposed for an eHealth solution aimed at enhanced pain management in general practice. The green area represents the patient perspectives and the blue represent the perspectives of the general practitioners. In the middle (area between green and blue) highlights the digital eHealth solutions generated by the matrix analysis. Abbreviations: e-Health: Electronic Health.

### A Pain Diary feature

Patients and GPs suggested that a pain journal could help patients to better communicate their pain to their physician, as well as helping patients to track and maintain an overview of their pain developments. Still one challenge was keeping journals in a language which was easy for patients and clinicians to understand.

### A tool for managing modifying processes

Some GPs suggested creating a feature which allowed patients to visualize and managing their biological, psychological, and social aspects and domains of their pain. This feature could empower GPs to gain a quick overview of the whole patient and identify processes which may influence the patient’s ability to respond to treatments.

### A Pain Education feature

Patients and GPs suggested that educating patients on their diagnosis, pain and pain management could help patients to better understand their pain and empower them to take charge of their conditions, which was a desired outcome of patients. Furthermore, gaining knowledge of the pain was believed to decrease worry and help patients communicate their pain.

### Personalised toolkit for management

Patients suggested having a feature with inspiration and different suggestions which patients could try out between consultations could help them to explore different approaches to managing their pain between consultations.

### Information features for relatives

Patients and GPs suggested how involving relatives could reduce patients’ feelings of stigma and guilt related to not being able to do what they used to and make it easier to express their pain and self-management needs.

### A Contact person or a guide

Finally, participants suggested incorporating a feature with a contact person where they could seek support when the pain became unmanageable, or someone (e.g., a person with the lived experience of pain) they could contact to ensure that they were on the right track with managing their pain.

## Discussion

### Principal Findings

This study integrated real-world insights from both patients and GPs and revealed distinct roles and challenges faced by each group in pain management. Patients grappled with understanding and coping with pain, impacting various aspects and domains of their lives, including daily activities, social interactions, and healthcare navigation. They expressed a need for a personalised, user-friendly platform featuring educational resources and pain management tools. Interestingly, GPs echo these sentiments, emphasizing the importance of individualization, strategic follow-up, and potential involvement in structured plans and increased information exchange during follow-ups. These findings significantly advance our understanding of designing eHealth solutions tailored for GPs and patients, effectively bridging gaps in chronic pain management. We identified where the communication gap exists, as well as the features and functionalities required to overcome this gap. This study expands our current understanding by introducing real-world key touchpoints and core features that have implications for future context-specific designs crucial for the successful implementation of eHealth solutions for pain management in general practice.

Our findings identified several challenges regarding pain management in clinical and in patients’ everyday settings, but also how this management spanned across multiple domains; patients and GPs had to master several tasks to ensure successful compliance to management ^12,50,74^. A key feature was how patients described that their pain was not solely a bio-medical phenomenon but also the degree pain impacted their social and work lives, as well as their mental health ^12^. While GPs recognized this and aspired to be supportive, our results indicated challenges in understanding patients’ situations, impeding effective collaboration. Johansen et al ^27^ demonstrated that a specific facet in management, such as patient education, extends across multiple domains and presents diverse perspectives within the context of pain management; often, the challenges and needs associated with these facets reveals itself in various dimensions and differ between healthcare practitioners and patients ^27^. Our findings provide novel insights into this, with both patients and GPs emphasizing the importance of patient education in eHealth solutions. However, patients were focused on the need for information to be short, concise, and understandable, possibly to prevent information overload and discontinuation of use, as described by Swar et al ^68^. GPs, on the other hand, were more focused on exploring eHealth tailoring to ensure that information was delivered within teachable moments ^42,43^ and that patients remained motivated to explore their own conditions, as described by Lusteria et al ^45^. This underscores the imperative of involving each patient in decision-making processes; failure to do so may lead to a mismatch between the available management options and the unique circumstances of each individual patient. By exploring the intersection between patient-GP perspectives and understanding these divergent dimensions further, future research can use strategies, such as participatory methods, to better support patients in navigating the healthcare system and with self-management of their pain ^61^.

The contrast between the design objectives (needs and challenges) generated in the matrix-analysis and the existing clinical guidelines for non-pharmacological management of chronic pain is evident ^29,44,57^. While clinical guidelines often emphasize generic non-pharmacological approaches, such as physical activity, education, and psychological interventions, our findings underscore a desire for more in-depth personalised approaches and features aimed at managing pain – from both sides in the clinical setting ^44,46^. Patients and GPs highlighted the importance of personalised and adaptable tools, including a pain diary, pain education feature, informational resources for relatives, and a designated contact person or guide. These tools aim to enhance communication, monitor the domains of living with pain, and facilitate learning about the condition to adjust to new self-management strategies. This contrast suggests a potential gap between the current guidelines and the more personalised needs expressed by those directly involved in real-world chronic pain management. Therefore, our findings may set the stage for new ways to integrate more personalised eHealth solutions and self-management recommendations that accommodate individual experiences and needs, while still ensuring alignment with evidence-based practices and clinical guidelines.

Healthcare is undergoing significant changes with increased digitization ^3,35,70^. These potential benefits require a careful approach and active user involvement in developing innovative solutions ^35,53,61^. Our findings identified core features for an eHealth solution supporting GPs’ or patients’ individual management, enhancing collaborative pain management. In our study, the analysis revealed the necessity for improved clinical communication between GPs and patients to support self-management and collaboration. This involves addressing temporal dimensions, monitoring progress over time, and tailoring tasks to individual needs during management ^19,30^. Patients faced difficulties effectively communicating their pain to GPs, while GPs described uncertainty in understanding the patients’ problems. Patients expressed a desire for control and decision-making autonomy but acknowledged the need for occasional guidance in their self-management. Conversely, GPs highlighted the complexity of patients’ pain experiences, emphasizing the time and effort required for effective engagement and role negotiation. The use of eHealth can facilitate patient-GP collaboration, supporting shared decision-making and helping GPs guide patients in managing their pain ^33,61^. Both patients and GPs noted the laborious and ongoing nature of pain management, aligning with findings by Corbin and Strauss, emphasizing the sustained effort required to address chronic pain’s multifaceted and temporal challenges ^12^. While literature highlights digital technologies’ potential to support collaboration in primary care ^1,53,65^, questions remain about how technology can facilitate shared vocabulary and negotiation between patients and GPs ^28^. This underscores the fragile foundation of eHealth, emphasizing the need for a continued engagement of stakeholders and involving them in decision-making to capture unique experiences, identities, and distinct needs ^58,67^.

While several trials have explored and documented the potential opportunities and benefits of incorporating various eHealth solutions in pain management, there are significant challenges in incorporating them into complex healthcare contexts ^15,17,23,52,63^. These challenges include limited participation from end-users, such as patients, and a lack of consideration for existing care processes ^24,54^. In our study, GPs and patients suggested a pain diary where patients could enter their symptoms via a specially customized features, such as their pain levels and processes as they emerged to foster acceptance and behavioral change. The individual needs are placing a demand on the healthcare system to engage in dialogues and assist patients in navigating their unique circumstances. These varied patient profiles, each with its specific needs, may necessitate additional proactive and innovative strategies that encourage collaboration between healthcare professionals and patients to effectively tackle these challenges. Importantly, while patients generally accept personalised self-tracking eHealth, there is also uncertainty about whether this leads to behavioural changes ^5,38,66^, with some studies indicating that self-reflection via quantified self may not induce behavioural change in adults with chronic conditions and, in some cases, may even cause harm ^36,38,45,66^. Essentially, these uncertainties necessitate future research to investigate and document the potential opportunities and downsides linked to the integration of eHealth solutions in pain management to ensure that the benefits surpass the potential harms.

### Strength and Limitations

The decision to exclusively include patients and GPs in our study may have limitations, as evidence suggests that incorporating the perspectives of relatives could offer a more comprehensive understanding of the challenges and dynamics in managing pain ^11^. Emphasizing the expansive scope of eHealth solutions, our findings are confined to visions applicable to general practices. Nevertheless, it is conceivable that these findings hold value for diverse settings, influencing the trajectory of patients across various sectors. Future studies should aim to understand how these findings may (not) align with design objectives that matter in other clinical settings. Although we identified several key features, these are overarching needs and are not ready for implementation. Therefore, future research should provide a more nuanced description of how these features, such as a pain diary, should manifest in practice. eHealth solutions, despite their potential benefits, face many challenges, such as maintaining sustained patient engagement; studies indicate that 97% of users discontinue their interaction with a mental health app within two weeks of downloading it ^6,8,62^. Future research addressing compliance processes that may lead to (dis)engagement and long-term commitment is needed to understand the processes influencing compliance, engagement, and long-term commitment of eHealth solutions. The strength of our study is in the thorough data analysis, where two independent members conducted coding in the data analysis. While a range of methods are applicable in studying various health conditions and eHealth adoption, workshops have several strengths, such as providing rich insights, fostering participant engagement, and allowing for in-depth exploration of complex phenomena ^16,60^. We encourage reflections on how the experiences interact with their interpretations, fostering a more comprehensive approach to eHealth integration in health-promoting initiatives.

### Conclusions

The findings reveal both contrasting and shared viewpoints on eHealth designs, providing insight into end-user perspectives for effective pain management. Our findings underscore the crucial role of effective clinical communication in understanding each patient’s overall clinical presentation, while shedding light on the significant challenges experienced by both patients and GPs in the clinical encounter. This novel knowledge offers promising opportunities for designing an eHealth solution to enhance personalized pain management in general practice.

## Data Availability

Data will be available upon reasonable request to corresponding author.

## Acknowledgments

The authors would like to thank Mads Christensen for his role in the execution of the two workshops.

## Notes

### Competing Interest Statement

All authors state no conflict of interest. MSH has received support from non-industrial professional, private, and scientific bodies (reimbursement of travel costs and speaker fees) for lectures on pain, and he receives book royalties from Gyldendal, Munksgaard Denmark, FADL, and Muusmann publications. Otherwise, none of the authors declare conflicts of interest.

### Funding Statement

This work is funded by the Foundation for General Practice (Danish: Fonden for Almen Praksis) and Nord-KAP-the Quality Unit for General Practice in The North Denmark Region (Danish: Kvalitetsenheden for Almen Praksis (Nord-KAP)). The funders had no role in the study design, data collection and analysis, decision to publish, or preparation of the manuscript.

### Author Declarations

This study was deemed exempt from ethical approval by The North Denmark Region Committee on Health Research Ethics due to the non-interventional nature of the study and the data that was collected. Signed informed consent was obtained from all participants. All data were stored on a secure fileshare.

