## Supplementary Material for "Supporting Self-management Through eHealth - Exploring the Needs, Challenges and Solutions in General Practice: A Qualitative and Participatory Design Study"

### Indhold

**Appendix 1 – Case vignette.**

Hanne is 42 years old and works as a healthcare assistant at a nursing home. Five years ago, she divorced her ex-husband and is now a single parent to her two children aged 10 and 12. Over the past three months, Hanne has experienced pain in the lower part of her back. The pain worsens when she is at work and when she does household chores at home, but it subsides during the weekends when she can relax a bit more. Hanne doesn't feel that her back pain has improved at all in the past three months. On the contrary, they have gradually worsened slightly, and over the past couple of weeks, Hanne has experienced that the pain has started to affect her everyday life more and more. Therefore, she consults her general practitioner. After a thorough examination, the general practitioner diagnoses "non-specific lower back pain" since he finds no signs of radiating symptoms or indications that Hanne's back pain could be due to another illness. There is therefore no indication for further investigation, and Hanne's doctor prescribes Ibuprofen and refers her to back training with a physiotherapist.

Months go by, and neither the back training with the physiotherapist nor the Ibuprofen has any particular effect. However, Hanne continues to take the pain-relieving medication as a precaution. Due to the lack of effect, Hanne again consults her general practitioner after 3 months. Hanne's doctor follows up on the previous examination, but the conclusion is the same: "non-specific lower back pain". The general practitioner refers her to a chiropractor instead. Following recommendations from family and friends, Hanne also chooses to seek treatment from a masseur and acupuncturist concurrently, but regardless of the approach she tries, all forms of treatment have only short-term effects.

Now, 9 months have passed, and it has become considerably harder for Hanne to go to work. In fact, within the past month, she has had to call in sick several days from work because the back pain has been so severe that she has been unable to handle her work tasks. The pain has also started to affect her mood and energy at home. She becomes irritated more easily. Moreover, Hanne often has to deprioritize regular chores at home and instead lie down due to the pain when she returns from work. The persistent pain greatly affects Hanne, and she becomes more and more resigned because the pain doesn't improve, no matter what she tries. The pain has also affected Hanne's sleep several times because before bedtime, she has started to speculate about what her future might look like.

**Note:** The case used during the workshops was originally in Danish, which was also the native language of the workshop participants.

**Appendix 2 - Inspiration Cards.**

A comprehensive list of themes for the inspiration cards.

**Situation cards: Situations/contexts**

- At work.
- During social events.
- At home.
- During one's leisure activities.
- At the doctor's office.
- At the physiotherapists.
- In the company of family.
- In the company of friends or colleagues.
- In the company of one's partner.
- Blank card: Write your/your own challenging situation here.

**Problem cards: Problems and challenges**

- Difficult to understand why I'm in pain.
- Don't know how I should manage the pain.
- Don't know how I should handle the situation.
- Fearful and worried about doing something wrong.
- The pain is unpredictable and erratic.
- Difficulty expressing myself to others.
- Feeling unseen, unheard, and misunderstood.
- Lack of support and understanding.
- Sleep problems.
- Blank card: Write your/your own experienced problems here.

**Inspiration cards: Suggestions for contents for an eHealth solution**

- Pain education - Learn to understand your pain.
- Learn to manage your pain.
- Express your pain to others.
- Serious illness - What signs should you be aware of?
- Learn to understand and navigate the healthcare system.
- Find your role in the treatment process.
- Prioritize and plan the use of your daily resources.
- Set meaningful goals and develop an action plan.
- Become more physically active in your daily life.
- Improve your sleep quality.
- Chat function for quick contact with healthcare professionals.
- Overcome negative thoughts and worries.
- Pain diary - Gain an overview of your pain.
- Blank card: Write your/your own solutions to the challenges here.

**Appendix 3 – Study procedure.**

| Duration/time | Agenda/plan | Responsibility |
|---|---|---|
| **Introduction**<br>15 min | Welcome and introduction to the research project (project aim, goal, agenda and ethical considerations).<br><br>Presentation of Smertevejleder (inspiration)<br>Participant information<br>Summary and questions<br>Informed consent<br>Presentation of the patient case | Lead researcher & facilitators<br><br>Facilitators |
| **Break**<br>5 min | Short coffee break. | |
| **Session 1: Identifying challenges**<br>45 mins | Introduction and instructions on how to complete the exercises.<br><br>Card exercises: Prompting question cards with ques to potential challenges during the course of treatment, and blank cards to note the participants perceived challenges. Post-its will be available for expanded descriptions of particularly relevant perspectives.<br><br>**Approach:**<br>• Participants read the case and questions aloud in the workgroups.<br>• By using the assignment cards, participants create a narrative outlining the journey of a CP patient (Blank cards fill gaps)<br>• Participants discuss potential challenges of either: 1) having CP and receiving treatment in general practice , or 2) managing patients with CP in general practice.<br>• Participants use the post-its to describe 'why' they see this as a problem (insights)<br><br>Insights are selected and presented to other groups (10 mins) | Facilitators |
| **Break**<br>5 min | Short coffee break. | |
| **Session 2: Identifying solutions.**<br>45 mins | Introduction and instructions on how to complete the exercises.<br><br>Card exercises: Prompting question cards with ques to e-health solutions during the course of treatment, and blank cards to note the participants ideas for potential solutions. Post-its will be available for expanded descriptions of particularly relevant perspectives.<br><br>**Approach:** | Facilitators |

| | | |
|---|---|---|
| | • Participants read the case and questions aloud in the workgroups.<br>• By using the assignment cards, participants create a narrative outlining the journey of a CP patient (Blank cards fill gaps)<br>• Participants discuss potential e-health solutions for either: 1) the challenges experienced as patient in general practice, or 2) the challenges experienced as a general practitioner managing people with CP in general practice.<br>• Participants use the post-its to describe 'why' they see this as a potential solution (insights).<br><br>Insights are selected and presented to other groups (10 mins) | |
| **Break**<br>5 min | Short coffee break. | |
| **Session 3: Prioritization and design specifications.**<br>45 min. | Introduction and instructions on how to complete the assignments of the final phase. The participants will be asked to choose the 1-2 key features for an e-health solution, which they perceive as being the most relevant ones.<br><br>• Participants are asked to choose two specific solutions, which they are asked to explore in depth (what, how, when, why, whom, etc.).<br>• Post-its are used for elaborating the participants perspectives.<br>• The session will be concluded with a presentation of the visions and the storyboards of each group in plenum and feedback.<br>• The facilitator notes visions and values on whiteboards as we go along. | Facilitator |
| **Summery and conclusion**<br>15 min. | Questions, practicalities and handing in permission to record data. | Facilitator and lead researcher. |
